# Estimating the false-negative test probability of SARS-CoV-2 by RT-PCR

**DOI:** 10.1101/2020.04.05.20053355

**Authors:** Paul S Wikramaratna, Robert S Paton, Mahan Ghafari, José Lourenço

## Abstract

**Introduction:** Reverse transcription-polymerase chain reaction (RT-PCR) assays are used to test for infection with the SARS-CoV-2 virus. RT-PCR tests are highly specific and the probability of false positives is low, but false negatives are possible depending on the individual, swab type and timing as infection progresses.

**Objectives:** To determine how the false negative test probability in infected patients depends on the time since symptom onset and swab type.

**Methods:** We use General Additive Mixed Models to analyse publicly available data from patients who received multiple RT-PCR tests and were identified as SARS-CoV-2 positive at least once.

**Results:** We identify that the probability of a positive test decreases with time after symptom onset, with oropharyngeal [OP]samples less likely to yield a positive result than nasopharyngeal [NP] ones. We also comment on the likely false negative rates in cohorts of patients who present for testing at different clinical stages and assess the robustness of these estimates to the probability of false positive tests.

**Conclusion:** NP samples are more sensitive than OP samples. The later an infected individual is tested after the onset of symptoms, the less likely they are to test positive. This has implications for identification of infected patients, contact tracing, and also for the discharge of convalescing patients who are potentially still infectious. There is an urgent need for data from asymptomatic and presymptomatic individuals.

## Introduction

Currently, most SARS-CoV-2 infected individuals are identified by successful amplification of the virus from throat or nasal swabs in the reverse transcriptase polymerase chain reaction (RT-PCR) assay. These tests are highly specific but there are many reasons why sensitivity is imperfect [1]. Indeed, multiple studies have observed negative RT-PCR results on at least 1 occasion for SARS-CoV-2 infected individuals [1–8]. Such false-negative results have implications for correct diagnosis [9] and subsequent community transmission [10], and thus for control initiatives, not only during emergence but also in any subsequent waves of transmission. For example, even in countries in Europe and Asia where outbreaks now seem to be under control, it remains unclear what fraction of the population has already been exposed and therefore the extent to which there remains the potential for large outbreaks following the reintroduction of the virus. Successful attempts to test and trace resurgent outbreaks will require a proper understanding of how test results alone cannot confirm or deny that an individual is infected with the virus, but also need to be taken in context with the other available information (such as the timing and type of test performed).

A series of previous studies have described cohorts of tested individuals. Ai and colleagues [2], for instance, retrospectively considered 1014 infected patients of whom 413 (41%) tested negative by RT-PCR at initial presentation. Xie et al. [1] similarly considered 167 infected patients of whom 5 (3%) tested negative by RT-PCR at initial presentation. Fang et al. [3] found that RT-PCR was only able to identify 36/51 (71%) of SARS-CoV-2 infected patients when using swabs taken 0-6 days after the onset of symptoms. Liu et al. [8]Zhao et al. [7] have demonstrated that the proportion of positive tests among infected patients dropped with each week since the onset of symptoms, and Luo et al. [11] reported that the initial sensitivity of throat swabs in secondary contacts was 71%. Meanwhile in a study of 213 patients, Yang et al. [4] found lower positive test rates from throat swabs (24%) compared to nasal swabs (57%).

Although these particular studies relate to longitudinal testing of infected patients, the data is not disaggregated per patient. Some authors have however presented sequential test data from individual patients [5,6,12]. Here we use the latter type of data to characterise how the probability of a false-negative test result depends on the number of days between the onset of symptoms and the performance of the test, and how this is affected by the site from which swabs are taken. We also estimate the number of false-negatives in different cohorts of tested individuals, under the assumption that they are only tested once; and assess the sensitivity of these results to the specificity of the test. Our results have implications for both existing estimates of SARS-CoV-2 prevalence and the likelihood of correctly identifying the infected status of an individual, where these rely solely on RT-PCR tests.

## Methods

### Data sources

Data for all sources were read directly from figures. To qualify for inclusion, studies had to report extractable results for longitudinal RT-PCR tests from individual patients who tested positive for SARS-CoV-2 at least once. The timing of swab collection had to be stated as the number of days after the patient became symptomatic. We identified seven studies that met these criteria and reported nasopharyngeal swab results [5,12,14–18] with two also reporting results for oropharyngeal swabs in the same patients [5,12]. Several studies with longitudinal patient data [6,19,20] only vaguely described the sampling location (e.g. “upper respiratory tract”) or did not make clear which results belonged to which patient. Other types of sample - such as sputum and saliva - were not considered as the collection methods were different to swabs. These 7 studies provided data from 786 tests on 95 patients for nasopharyngeal and oropharyngeal swabs. Summary information about the included studies is presented in Table 1.

**Table 1:** Summary of the data sources used in this study. Where available, the target gene for the PCR is noted, as is the definition of an infection for an individual study. Counts of cases and swabs (nasopharyngeal, NP, and oropharyngeal, OP) are also reported. * Used in this study [available], ^1^ The study used a combination of nasopharyngeal and mid-turbinate swabs.

| Paper | Target gene | Swab type(s) | Infection definition and case description | Number of cases* | Number of swabs |
| --- | --- | --- | --- | --- | --- |
| Danis et al. | Not specified | Nasopharyngeal | All patients were isolated in hospital and were RT-PCR confirmed. 5 of the 6 presented symptoms. One patient was only confirmed using endotracheal aspirate samples and was excluded from the analysis. | 4 [6] | 38[NP] |
| Kujawski et al. | 3 targets on N | Nasopharyngeal and oropharyngeal | 7 of 12 patients were hospitalized with 5 remaining at home. All were RT-PCR confirmed. One patient was excluded due to a poorly defined symptom start date. | 11 [12] | 86[NP] 90[OP] |
| Lescure et al. | RdRp-IP1 and RdRp | Nasopharyngeal | All 5 patients were admitted to hospital and were RT-PCR confirmed. | 5 [5] | 42 [NP] |
| Seah et al. | Not specified | Nasopharyngeal | All 17 patients were recruited while in hospital and were RT-PCR confirmed. | 17 [17] | 132 [NP] |
| Wyllie et al. | Multiple targets on N (CDC assay) | Nasopharyngeal | All patients were RT-PCR hospital patients with symptoms. Of the 44 patients, 22 had longitudinal NP testing time series. | 22 [44] | 53 [NP] |
| Young et al. | N, S and ORFab1 | Nasopharyngeal and oropharyngeal | All patients were admitted to hospital and RT-PCR confirmed. | 18 [18] | 230 [NP] |
| Zou et al. | N and ORF1b | Nasopharyngeal<sup>1</sup> and oropharyngeal | Cases are referred to as patients but hospitalization is not explicitly stated. All cases are RT-PCR confirmed. One patient was asymptomatic. | 17 [18] | 61 [NP] 55 [OP] |
|  |  |  |  | 95 [120] | 642 [NP] 145 [OP] |

### Data analysis: Estimating RT-PCR sensitivity

Data were analysed using binomially-distributed (logit-link) generalised additive mixed models (GAMM) with the package mgcv [21] in the statistical software R version 3.6.3 [22]. We tested hypotheses that the probability of a positive result will change through time after symptom onset, that different swab type may have different detection probabilities and that each study may have a different baseline detection probability (due to, for example, differing testing procedures). The effect of the number of days since symptom onset was modelled as a continuous smooth function (cubic regression splines), while swab location and data source were included as two-level categorical variables. Random effects were included in the form of patient-specific smooth functions, modelling between-patient differences in the probability of returning a positive test through time. All of the models we examine included this random effect, as patient samples were pseudo-replicated by design. Models were compared in a stepwise down procedure from the most complex structure using Akaike Information Criterion (AIC). The difference in AIC values (ΔAIC) values were calculated in relation to the lowest AIC value. Detailed methodology is available in Supplementary File 1.

### Data analysis: Estimating the number of false-negatives in a cohort of tested individuals

Results from Bi et al. [13] suggests that the probability of an infected individual getting a positive RT-PCR test of SARS-CoV-2 after a given number of days since the onset of symptoms follows a gamma distribution with shape 2.12 and rate 0.39 (see Figure 3 and Table S2 in [13]). We can use this together with our results on RT-PCR sensitivity and apply Bayes’ Theorem to recover the distribution of the time from onset of symptoms to getting tested (see Supplementary File 1l), which ends up as a distribution with a heavier tail (than the original gamma distribution) because the false-negative test probability increases with time.

**Figure 1:**
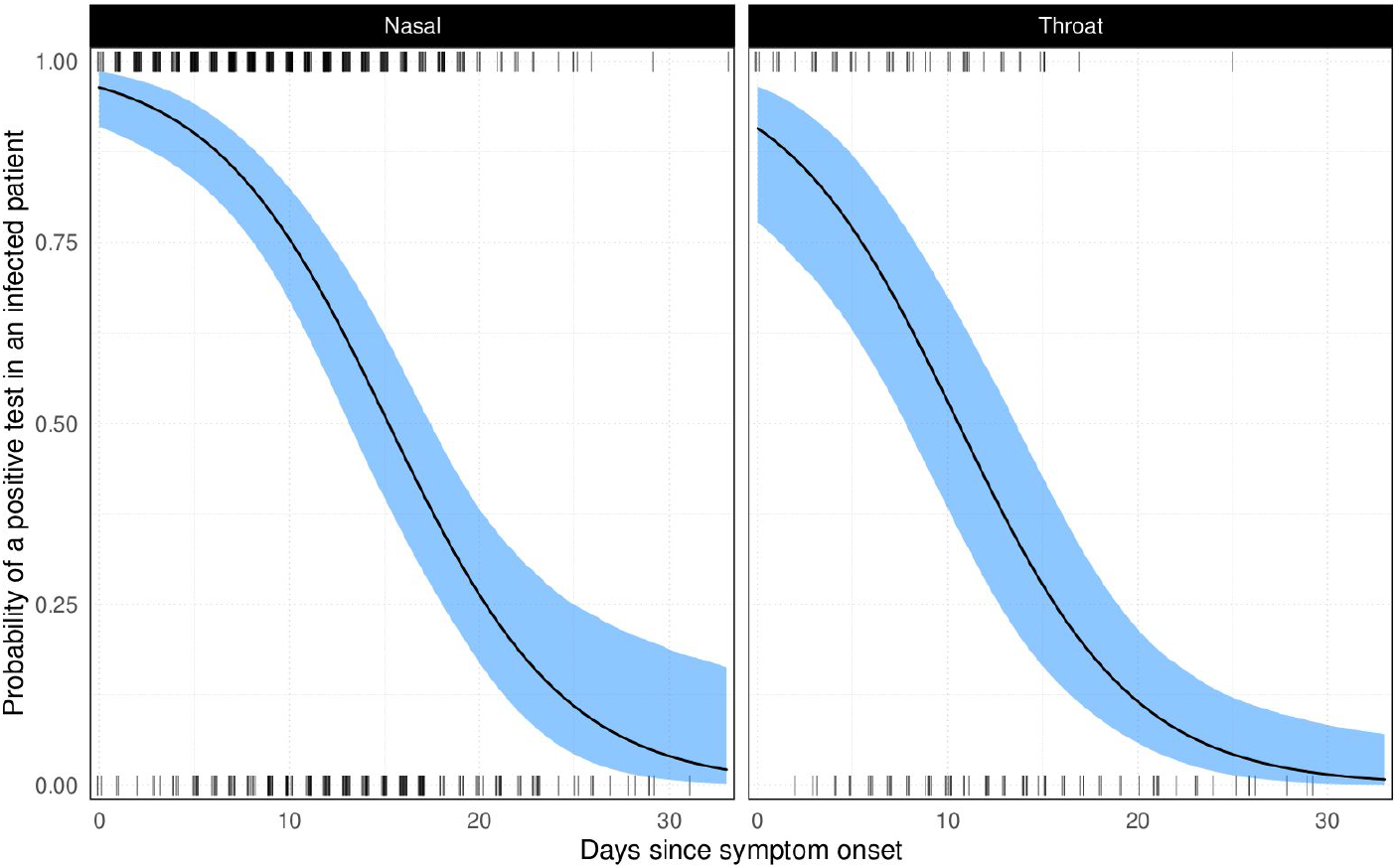
Model fit of a binomially-distributed GAMM to longitudinal RT-PCR test data. Tick marks denote positive (top) and negative (bottom) tests (jittered on the x-axis for visual purposes). The black line shows the model fit and blue ribbons the 95% confidence intervals on the fixed effects. The left-hand panel gives results for nasal swabs and the right throat swabs.

**Figure 2:**
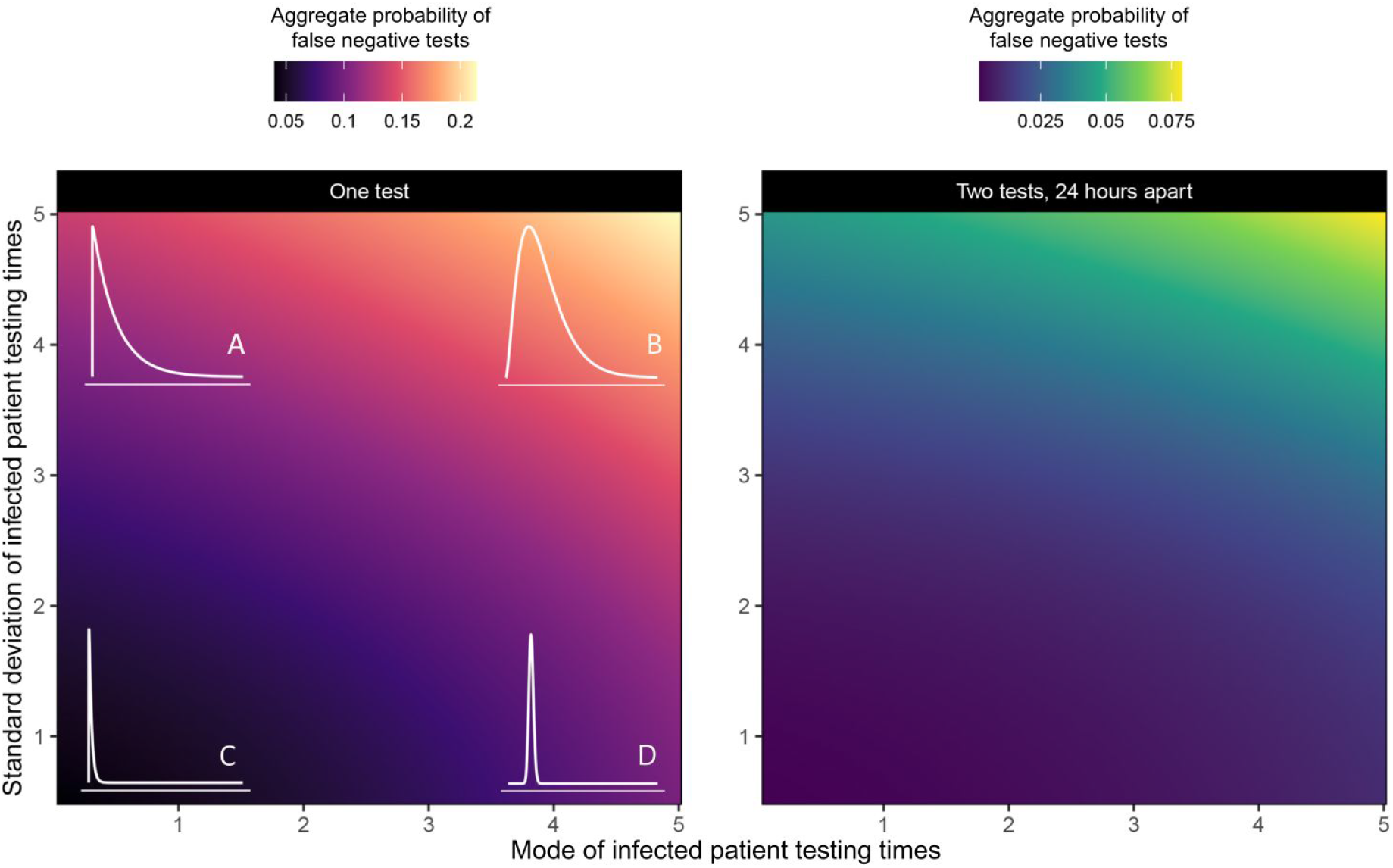
Surfaces showing the aggregate probability of false negative tests (denoted by colour) for Gamma-distributed cohorts of people being tested for SARS-CoV-2 infection. Each point on the surface is a result for a unique parameterisation of the distribution. The x-axis shows the mode of the testing time Gamma distribution and the y-axis the standard deviation. The left-hand panel shows the error rate for one test, while the second panel shows the error rate for two tests taken 24 hours apart. Illustrative distributions are drawn in the corresponding corners of the first panel (these are the same in the second panel), with subplots **A**-**D** showing illustrative examples at the 4 extremes. **A** gives a scenario where most patients are tested early but with a “long-tail” of patients taking a long time to be tested. **B** shows a scenario where patients are mainly tested later, with a similarly long tail. Scenarios **C** and **D** represent scenarios where patients are consistently tested early or late (with very thin tails).

**Figure 3:**
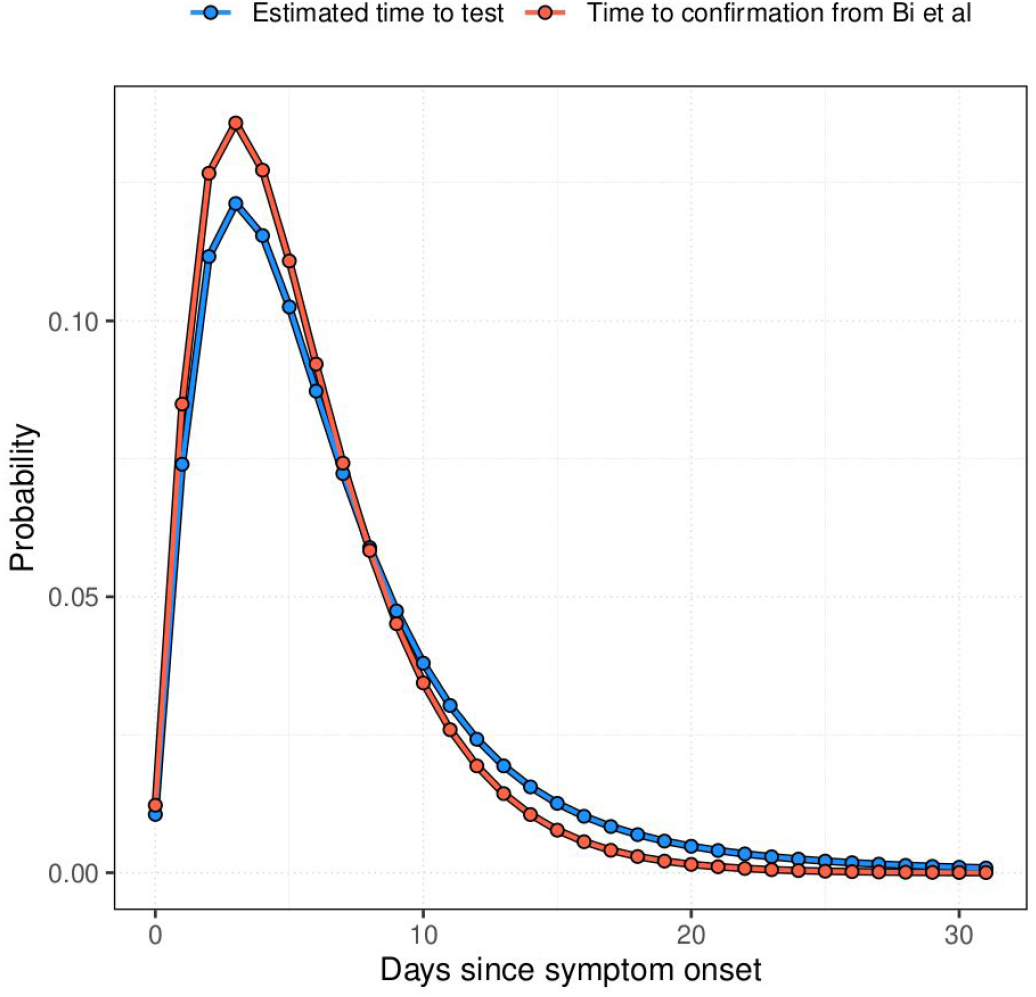
Comparison of the discretised distributions of time to confirmation among symptomatic individuals. From Bi et al. and the distribution of time to test that we estimate, combining this with our estimate of the false-negative test probability from nasal swabs, assuming false-positive tests are impossible (see Supplementary File 1).

If we assume further that this distribution is generally representative and does not vary over the course of the epidemic, we can use it to estimate how many infected individuals are incorrectly identified as uninfected among a group of symptomatic tested individuals who are only tested once. We can further explore how the false-positive test probability affects these estimates and illustrate this using the numbers of tests performed and positive test results from the UK and South Korea as of 20th March 2020 (UK: 5.1% positive [3277 / 64621 positive tests]; SK: 2.7% [8652 / 316664 positive tests] [16]). It is important to stress that this exercise is illustrative rather than assertive: we show how accounting for the false-negative and false-positive test probabilities affects the estimate of the number of infected individuals among those tested, taking broad assumptions (e.g. all individuals only tested once; that the distribution to test is as we have assumed; that all those tested are symptomatic) any or all of which is likely to be violated in these datasets. Therefore, we do not make specific predictions but are rather presenting a sensitivity of scale for the overall impact of accounting for the false-negative and false-positive test probabilities.

### Ethical statement

No ethical approval was required for this meta-analysis of publicly available data.

### Reproducibility

To ease reproducibility and allow reuse of our findings, we provide the main results, R scripts and data used as Supplementary Files 2, 3 and 4 respectively.

## Results

Our most complex model included two fixed effects (a smooth effect of day and a different intercept for each swab type) as well as a random intercept for each study and each patient (model AIC = 805.79). Removing the random intercept for study was supported (AIC = 805.31, ΔAIC = −0.47), suggesting that the baseline probability of detection was consistent across different cohorts, test targets and sample processing procedures. Excluding swab type was not supported (AIC = 813.15, ΔAIC = 7.83), nor was excluding the effect of days since symptom onset (AIC = 974.30, ΔAIC = 168.99). The final model structure with the highest support contained the fixed effect of swab type, the effect of time since symptom onset and a random intercept for each patient. The full model output is given in the Supplementary File 1.

Swabs taken from the throat immediately upon symptom onset were predicted to be 5.64% less likely to yield a positive result than a nasal swab (logit-scale effect size −1.00 CI [−1.54, −0.46]). The probability of a positive test decreases with the number of days past symptom onset: for a nasal swab, the percentage chance of a positive test declines from 96.40% [90.98, 98.6] on day 0 to 75.47% [66.88, 82.51] by day 10 and only a 3.30% [0.53, 17.90] chance of a positive result by day 31; for throat swabs the probabilities were 90.76% [77.84, 96.52], 53.00% [38.27, 67.46] and 1.23% [0.18, 7.86] for day 0, 10 and 31 respectively. The model fit is shown in Figure 1 and the underlying quantitative results (data) are available in Supplementary File 2.

### Visualising the impact of time to test on false-negative test probabilities

As shown above, the probability of a false-negative test result depends on the number of days since symptom onset. This means that simple reports of positive and negative test counts among individuals who are only tested once will underestimate the true number of positive tests in a cohort. We can illustrate the potential impact this has on average false-negative test probabilities by supposing that the time from onset of symptoms to testing follows a gamma-distribution.

Figure 2 explores how varying the shape and rate of this distribution affects the average false-negative test probability in a cohort, and highlights that in scenarios where infected individuals are typically tested late, the false-negative test probability can be 4 times larger than when patients are typically tested early. We also show how the probability of incorrectly identifying an individual as uninfected due to a false-negative test considerably reduces if all negative tests are repeated 24 hours later. We note that the realised error rate (the actual proportion of false negative tests) will be proportional to the underlying prevalence of infection; only if everyone in the cohort is infected would the probability of a false negative equal the proportion of negative tests (as there will be no true negatives from uninfected individuals).

### Estimating the number of false-negatives in a cohort of tested individuals

We further demonstrate how the results of Figure 2, together with assumptions about the false-positive test probability, might affect testing outcomes in practice.

Figure 4 shows that when the false-positive test probability is very small then the estimated number of infections among those tested is increased by around 30%, but this estimate decreases linearly as the false-positive test probability increases. In fact, for some critical (yet small) value for the false-positive test probability, there will be more false-positives than false-negatives. Moreover, the false positive test probability has a bigger impact when the percentage of the original tests that were positive is smaller (this follows directly from the underlying derivations - see Supplementary Methods in Supplementary File 1).

**Figure 4:**
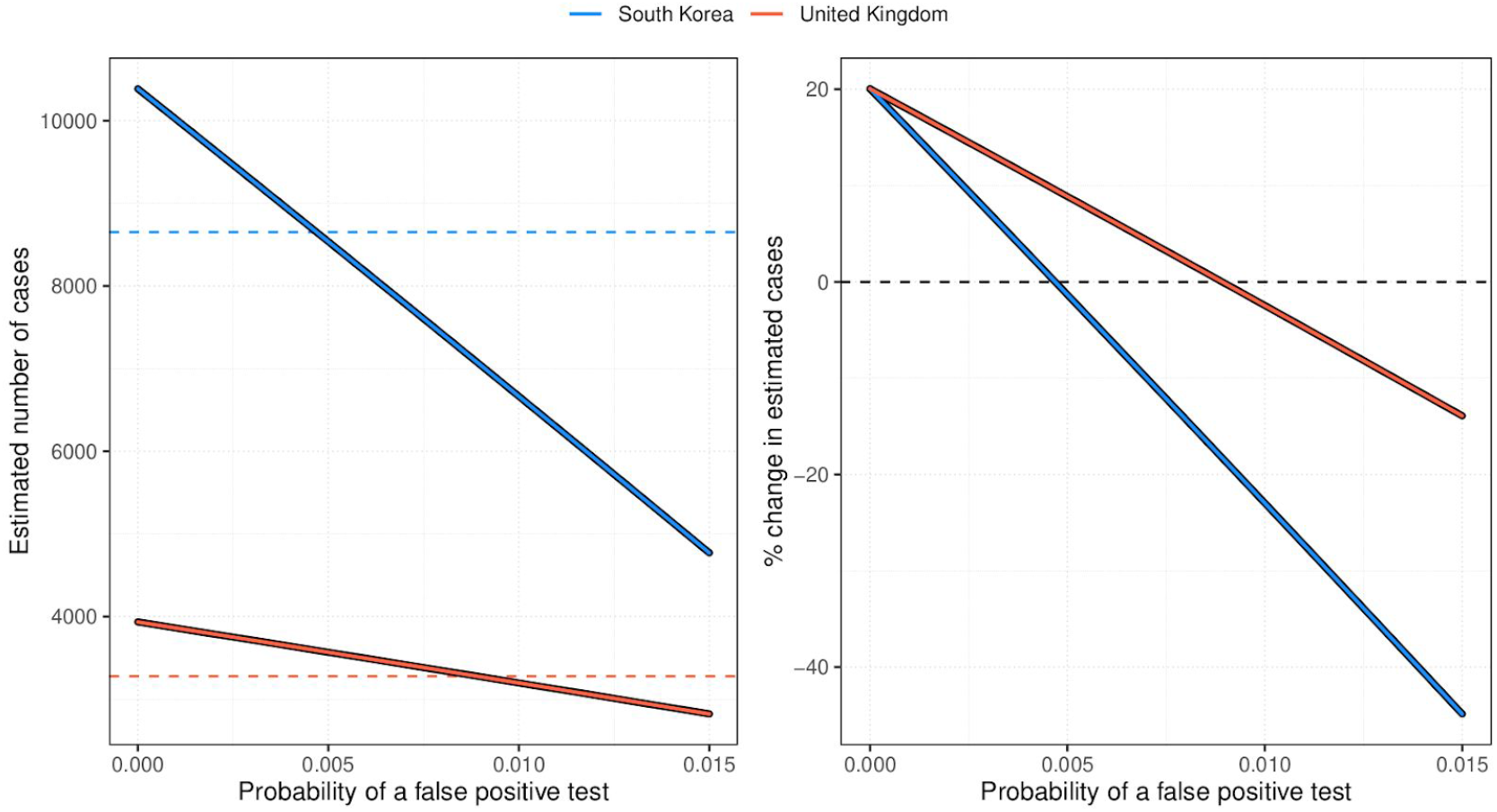
Impact of false-negative and false-positive test probabilities on estimated number of infections. **(A)** Impact of the false-positive and false-negative test probability on the estimated total number of infected individuals among those tested in South Korea (red) and the UK (blue); dashed line corresponds to number of positive tests conducted as of 20th March 2020. **(B)** Similar, but now showing the percentage change from the number of positive tests to the estimated number of cases.

Overall this illustrates 3 important things: that for a zero or very small false-positive test probability, the true number of infected individuals among those tested will be substantially larger than the number of positive tests; that increasing false-positive test probabilities decrease these estimates until they eventually go negative (even for quite small values of the false-positive test probability); and that such decreases are more severe in situations where the apparent prevalence among those tested is lower.

## Discussion

Testing from single throat or nasal swab by RT-PCR is not guaranteed to yield a (true) positive result for SARS-CoV-2 infection and this probability decreases with time since the onset of symptoms. In other words, the longer the time from the onset of symptoms until a suspected case is tested, the more likely a false-negative result. Repeat testing of suspected but RT-PCR negative infections may not always be feasible, but our results suggest that repeat testing drastically decreases the chances of failing to identify infected individuals.

Meanwhile, failing to account for the possibility of false-negative tests potentially biases upwards many of the existing estimates for case and infection fatality rates of SARS-CoV-2 e.g. where they rely on perfect sensitivity among international travellers [24,25].

We also show how even small false-positive test probabilities can have an opposite impact on any assessment of the “true” number of infections in a tested cohort and hence bias case and infection fatality risk estimates in the opposite direction. Better understanding of the false-positive test probability and accounting for precisely when and how individuals have been tested would therefore improve the quality of any estimates that rely on the number of positive tests in a cohort of tested individuals.

Our results have important implications for SARS-CoV-2 testing strategies. Presently, RT-PCR testing regimes vary significantly between countries, determined both by policy decisions, testing capacity and perceived incidence. Some countries opt (or, rather, are able) to test large portions of the population, including those who are asymptomatic or self-isolating with mild symptoms. In countries such as South Korea, where testing has been thorough [26,27], the distribution of test timing is crucial: if many of those tested were infected some time ago but only had mild or asymptomatic infections (and therefore did not present for treatment), they will be more likely to return a false negative result. In countries that do not currently have mass testing, there are calls for testing to be expanded to the population at large with the aim of determining how many people carry, or have recently carried the virus. While RT-PCR testing of key workers is of great importance (particularly those working with vulnerable groups), our results suggest that there may be little benefit to testing indiscriminately; in fact, conducting a single test on someone who had symptoms 10 days ago will have a nearly 25% chance of being a false negative (using a nasal swab; 47% for a throat swab). As a means of determining population level exposure to SARS-CoV-2, serological tests of known specificity and sensitivity are far more likely to provide an accurate profile (if these are known, then recovery of the likely level of population exposure is possible, even if it is not possible to determine which individuals have been exposed. This, however, would assume that all those infected will go on to develop and maintain detectable serological responses, which may not always be the case in e.g. mild infections, or across all age groups [28–33]).

In almost all countries, tests will be conducted on patients presenting with symptoms at a hospital in order to streamline treatment and prevent further infection. We do not suggest that the problem of false negatives is under appreciated by medical professionals; it is presently recognised by both the guidelines from the World Health Organisation (WHO) [34] and the European Centre for Disease Control (ECDC) [35] that a single negative test is insufficient to rule out infection, with discharge criteria stating that a patient should only be released if two repeat tests return negative results. Early in the outbreak, clinicians used chest CT to look for evidence of SARS-CoV-2 in symptomatic patients who returned a negative result, minimising the risk of false negatives [2,36].

We also note that RT-PCR tests will return positive results even if the virus is inert - only by culturing the sample is it possible to verify that a patient is actually infectious. To date, the available information suggests that it is possible for individuals with mild disease to shed for at least 18 days [37] and those with severe disease for at least 20 [19]; but evidence is mounting that successful culture of virus is associated with high viral loads and perhaps also not yet having a detectable serological response [6,19,38]. Further characterisation of these important relationships is required, but it suggests that quantitative estimates of (still detectable) viral load and/or seroconversion could be used as tools to safely discharge infected individuals from quarantine or hospital [19]. In light of this evidence, our study cannot readily assist with such decisions not least because the higher false-negative test probabilities we report with time since symptom onset are themselves almost certainly related to lower viral loads with time; what would be more useful would be quantitative estimates of how viral load changes with time since symptom onset. Repeat measurements and calculation of viral load in infected individuals would allow more precise interpretation of RT-PCR results, specifically whether a “negative” or high cycle threshold (CT) result is consistent with the previous trend in that patient, or is anomalous, and so determine how likely the individual is to still be infectious. Modelling efforts could help enormously on this topic, but in the meantime our results suggest that multiple negative tests would be consistent with the viral load having reduced to a sub-infectious level, but a single negative test may only represent a spurious test result.

Major implications of our results relate directly to contact tracing of infected individuals. Since it is possible for infected individuals to test negative for infection even soon after the development of symptoms, this means that a single negative test alone is also insufficient for ruling out infection in their recent contacts, who may have already been infected and be currently in the latent phase or a- or pre-symptomatic themselves. It may therefore be prudent to assume that contacts of suspected infected individuals may be infected themselves and isolate and regularly test them until either there is firm evidence that the putative index case was not actually infected (or at least infectious) at the time of the contact, or assess if the incubation period for the contact has passed without evidence that the contacts have themselves been infected. Doing so would enhance infection control and also ultimately provide more detail on test sensitivity at all possible stages of infection.

In conclusion, we demonstrate how the sensitivity of the RT-PCR assay for detecting SARS-CoV-2 infection depends on the time from the onset of symptoms in symptomatic individuals, and show how nasal swabs appear more sensitive than throat swabs. In the absence of other testing procedures, this has implications for clinical decisions about treatment, and control / contact tracing decisions about who needs to be quarantined or can be released safely into the community. We also illustrate how, assuming that the false positive test probability is negligible, the count of positive tests underestimates the count of infected individuals in a cohort of tested individuals, which in turn has implications for estimates of case and infection fatality rates in the wider population. However, if the false-positive test probability is non-zero, then values as low as 0.5% - 1% could mean that the true prevalence among those tested is lower than suggested by the naive count of positive tests.

### Limitations

First, more data exists than we have been able to analyse. Many of the studies cited here (& others - e.g. [39]) have longitudinal data from more patients but which is not currently publicly available and we were unable to obtain, or is not disaggregated by swab type. Inclusion of this data would provide superior estimates, in particular if it is disaggregated into tests done from different samples via different routes in the same patient. Moreover, explicit reporting of dates when tests are performed in all patients (and not just those who test positive) would be especially useful to any subsequent similar analyses for SARS-CoV-2 or other emerging viruses. We thus advocate for such data to be made available more readily in publications and preprints.

Second, we have attempted to account for possible differences among labs performing RT-PCR tests and although we do not find any evidence in favour of this being relevant, nor is there enough evidence to rule it out. There may also be variation in terms of the gene that is targeted, or method of RT-PCR performed, which we have also not been able to consider due to lack of available data (e.g. some target / assay combinations may be more sensitive than others [40,41]). We have also assumed that all the patients have been correctly identified as infected in each study and that the specificity of the test is perfect; it is likely to be extremely high but perhaps not 100% and may similarly vary with assay type [42].

Third, we have attempted to account for possible differences among patients in their sensitivity to the test. In reality, one might expect this to be related to either the underlying severity of the infection (perhaps a higher chance of detection when disease is more severe) or, at least, viral load (higher chance of detection with higher viral load); neither of which we have been able to assess with currently available data. Furthermore, the data we use is from symptomatic patients, most of which were sick enough to require hospital treatment, and it is possible that RT-PCR is less sensitive in asymptomatic individuals (not least because there is no onset of symptoms and it is therefore unclear from which baseline test sensitivity should be measured) or those with more mild disease. A recent Italian study offered evidence that, among those testing positive, viral loads were equivalent in symptomatic and asymptomatic individuals [43]. This does not show, however, that viral loads are the same in both groups, but that they are equivalent conditional on a positive test, which is what we might expect if the probability of a positive test is indeed linked to viral load. If this is true, then it could be that many asymptomatically infected individuals are asymptomatic because their immune responses keep viral replication in check early on and so viral loads sufficient to result in a positive test may not be achieved. However, such a scenario might be difficult to square with the apparent transmission potential of asymptomatic individuals [44]. Better understanding of the sensitivity of the test in asymptomatic individuals is of paramount importance but was impossible to assess with currently available data.

Fourth, and related, we do not have access to testing data from the pre-symptomatic period of infection. The high detection probability at symptom onset suggests that viral load could be high during the incubation period, particularly just before the onset of symptoms. We would also expect that the chances of detecting infection on the day of exposure is next to zero, but the detection probability between exposure and prior to symptom onset will likely be contingent on a number of factors, including the initial infectious dose of SARS-CoV-2 and the length of the incubation period. More data is urgently needed to answer these questions, particularly given the estimated transmission potential in this period [39,45]

Finally, when estimating the true number of positives in a cohort of tested individuals we have to additionally assume that the distribution of the time to test is the same as we infer from our results here and the distribution of time to confirmation in Guangdong ([13]). Even if this distribution is broadly representative from country to country, it may not be consistent over time. For example, as testing capacity gets stretched, the time to test may increase and so too the probability of a consequent false-negative (or vice versa as testing capacity is enhanced). These particular results should therefore be taken as indicative rather than authoritative. Furthermore, these results only relate to the cohort of tested individuals rather than the population at large: they say nothing about the prevalence of the virus among those not tested. That said, individual hospitals, testing centres or studies will know the timings of their tests and can use such information in conjunction with the quantitative findings that we make available in Supplementary Files to assess how likely any one test is to represent a false-negative.

This work has advanced our understanding of the significant effect that false negative RT-PCR tests can have on the identification of SARS-CoV-2 infected individuals. We have demonstrated the sensitivity of population prevalence estimates to erroneous test results, and caution that single negative tests should not be overinterpreted. This is particularly pertinent when tests are used to determine if healthcare staff and carers are safe to work with those most vulnerable to disease, or whether to isolate contacts of suspected infections. As nations begin lifting strict social distancing measures and returning to some semblance of normality after the first epidemic wave, being able to dependably contact trace and test new infections will be critical to prevent resurgence. This work suggests caution should always be used when interpreting a single, SARS-CoV-2 RT-PCR test result.

## Data Availability

All the data used in this manuscript was already in the public domain.

## Conflict of interest

None declared

## Funding statement

MG was funded by the Biotechnology and Biological Science Research Council (BBSCR) grant number BB/M011224/1 and the Oxford-Radcliffe graduate scholarship from University College, Oxford. JL was supported by a Lectureship from the Department of Zoology, University of Oxford.

## Author’s contributions

PSW designed research; RSP, PSW, MG carried out research; all authors analysed results and wrote the paper.

## Supplementary Files

Supplementary File 1: detailed methods and R summary model output.

Supplementary File 2: comma-separated file with model output for Figure 1

Supplementary File 3: R script

Supplementary File 4: comma-separated file with data used

## Notes

### Competing Interest Statement

The authors have declared no competing interest.

### Funding Statement

No particular funding supported this research.

### Summary of Updates

Update with changes made post revision for Eurosurveillance.

