## Supplementary File 1 for "Estimating the false-negative test probability of SARS-CoV-2 by RT-PCR"

### Detailed methods and R summary model output

#### GAMM models

##### Summary model output

This is a representation of the fitted values in the final model for test sensitivity as returned by mgcv::summary in R:

| *Predictors* | *Estimate* | *SE* | *p* | *df (edf for smooth)* |
|---|---|---|---|---|
| Intercept (inc. Nasal swab) | 0.87 | 0.19 | **<0.001** | - |
| Swab type (Throat) | -1.00 | 0.28 | **0.0002** | 1 |
| Smooth term (Days since symptom onset) | - | | **<0.001** | 1.06 |
| RE Smooth term (Patient) | - | | **<0.001** | 63.13 |

| | |
|---|---|
| $R^2$ adj | 36.20 % |
| AIC | 805.31 |

##### Sensitivity of Zou et al estimates

We utilise data from Zou et al. (2020) who use a combination of mid-turbinate and nasopharyngeal swabs to constitute nasal samples. To determine if there is an effect of using this combination of different swab types on results, we coded the "swab type" variable to have a separate level corresponding to the nasal samples for Zou et al, then compared it to the best fitting model with only two levels in the swab type variable (AIC = 805.31). The inclusion of a Zou-specific correction was not supported (AIC = 805.81, ΔAIC = 0.50).

##### Estimating the false-negative error rate in cohorts of tested individuals

Using the GAMM model, we estimated the aggregate false negative rate for hypothetical cohorts of tested patients. To do this, we considered a range of Gamma distributions as parameterised by the mode and standard deviation. These distributions were used to describe the time between the onset of symptoms and patients being tested. The shape (S) and rate (R) parameters were written as functions of the mode (M) and standard deviation ($\sigma$) [23]:

$$R = \frac{M + \sqrt{(M^2 + 4\sigma^2)}}{2\sigma^2}$$

$$S = 1 + MR$$

We explored arrival time distributions with modes ranging from 0.1 to 5 days and standard deviations ranging from 0.5 to 5. We discretised the arrival time distribution ($\Gamma(x)$) to give the proportion of patients in a cohort being tested on a given day. These fractions were then multiplied by the estimated probability of a false negative predicted by the GAMM function (f(x)) for a single nasal swab on that day; summing these together gave the aggregate false negative rate (*P(Neg|Inf)*) for cohorts tested according to this particular arrival time distribution. To get the probability of 2 false-negatives 1 day apart, we simply took the product f(x).f(x+1) and used this in place of f(x).

#### Estimating the time to test

Let

- $\tau_i$ correspond to being tested on day i
- $\psi$ correspond to having a positive test result
- $\eta$ correspond to being infected

Then

$$P(\tau_i \cap \eta) = \frac{P(\tau_i \cap \eta \mid \psi) \times P(\psi)}{P(\psi \mid \tau_i \cap \eta)}$$

We assumed the test has perfect sensitivity, such that $P(\tau_i \cap \eta \mid \psi) \equiv P(\tau_i \mid \psi)$ since all individuals with positive tests must be infected, and so we estimated this for each day using the distribution of time to positive test results for symptomatic individuals from Bi et al. [13] (a gamma distribution with shape 2.12 and rate 0.39). We discretised this distribution (such that [0, 0.5) corresponds to 0 days from symptom onset, [0.5, 1.5) corresponds to 1 day after symptom onset etc) and truncated it to 31 days, which is the maximum number of days from symptom onset present in the data we analysed. This truncation has no practical impact because > 99.99% of the density of this particular gamma distribution is accounted for at this point.

Meanwhile $P(\psi \mid \tau_i \cap \eta)$ is the probability of a positive test result for infected individuals given the day of the test, which is exactly what we estimated in this study. Of course, $P(\psi)$ is unknown. This gives us

$P(\tau_i \cap \eta)$ but as we assumed that individuals are tested only once then $\tau_i \cap \tau_j = \{\varnothing\}$ for $i \neq j$ which means that we can easily retrieve:

$$P(\tau_i) = P(\tau_i \cap \eta) \,/\, \sum_j P(\tau_j \cap \eta)$$

and then the unknown $P(\psi)$ appears in every term on the RHS and so vanishes.

##### Estimating the true prevalence in a cohort of tested individuals

Supposing that all tests were performed the same number of days after symptom onset; we defined:

- $\alpha$ as the (unknown) true prevalence among those tested
- $\beta$ as the false-positive rate i.e. P(positive test | uninfected)
- $\gamma$ as the false-negative rate for tests done on that day i.e. P(negative test | infected)
- *T* is the total number of tests done on that day, of which a fraction q are positive

Then the true prevalence among those tested for infection is equal to the sum of (a) P(infected|positive test) multiplied by the number of positive tests and (b) P(infected| negative test) multiplied by the number of negative tests (i.e sum of the true positives and false negatives). These conditional probabilities can be separately rearranged via Bayes' Theorem and then added together to give:

$$\alpha T = qT \frac{\alpha(1-\gamma)}{\alpha(1-\gamma) + (1-\alpha)\beta} + (1-q)T \frac{\alpha\gamma}{\alpha + (1-\alpha)(1-\beta)}$$

When rearranging, this as a quadratic in $\alpha$ then we discover it has 2 roots:

$$\alpha = \frac{q - \beta}{1 - \gamma - \beta}$$

$$\alpha = 1$$

And so the first root allows us to estimate the true prevalence among the test cohort, while accounting for the false-negative test probability for those tested on that day.

In reality, however, individuals are tested on different days on which the false negative test probability depends, which makes it much harder to estimate $\alpha$ in this way. One way it can be done is to use the distribution for time to test to calculate the average false-negative test probability across all tests conducted, again assuming that all tests are done by nasal swab - here this gives a false-negative test probability of 16.71%. If we do this, then we can still apply the same equations as above and explore how accounting for the false-negative and false-positive test probabilities affects the consequent estimates of the true prevalence among those tested, which we illustrate for some different scenarios in the main text.

Importantly, this only tells us about prevalence in the test cohort and not in the wider population i.e. this does nothing to correct for not finding and not testing mild/asymptomatic cases (as discussed in the main text).
